# Detection Of SARS-COV-2 Variants Of Concern In Wastewater Of Leuven, Belgium

**DOI:** 10.1101/2022.05.12.22274823

**Authors:** Mandy Bloemen, Leen Delang, Annabel Rector, Joren Raymenants, Jonathan Thibaut, Bram Pussig, Lore Fondu, Bert Aertgeerts, Marc Van Ranst, Chris Van Geet, Jef Arnout, Elke Wollants

## Abstract

To investigate whether wastewater surveillance can be used as an early warning system to detect a rise in SARS-CoV-2 positive cases, and to follow the circulation of specific variants of concern (VOC) in particular geographical areas, wastewater samples were collected from local neighborhood sewers and from a large regional wastewater treatment plant (WWTP) in the area of Leuven, Belgium. In two residential sampling sites, a rise in viral SARS-CoV-2 copies in wastewater preceded the peaks in positive cases. In the WWTP, peaks in the wastewater viral load were seen simultaneous with the waves op positive cases caused by the original Wuhan SARS-CoV-2 strain, the Alpha variant and the Delta variant. For the Omicron BA.1 variant associated wave, the viral load in wastewater increased to a lesser degree, and much later than the increase in positive cases, which could be attributed to a lower level of fecal excretion, as measured in hospitalized patients. Circulation of SARS-CoV-2 VOCs (Alpha, Delta and Omicron) could be detected based on the presence of specific key mutations. The shift in variants was noticeable in the wastewater, with key mutations of two different variants being present simultaneously during the transition period.

We found that wastewater based surveillance is a sensitive tool to monitor SARS-CoV-2 circulation levels and VOCs in larger regions. This can prove to be highly valuable in times of reducing testing capacity. Differences in excretion levels of various SARS-CoV-2 variants should however be taken into account when using wastewater surveillance to monitor SARS-CoV-2 circulation levels in the population.

## INTRODUCTION

Severe Acute Respiratory Syndrome Coronavirus 2 (SARS-CoV-2) rapidly spread around the globe following its emergence in China in December 2019. The virus has infected more than 306 million people worldwide of which more than 6 million died (data of April 1^st^ 2022) [1]. Rapid identification of infections is needed to detect and manage SARS-CoV-2 transmission in communities. To this end, SARS-CoV-2 surveillance has mainly focused on the identification of infected individuals with symptoms. However, this approach will not identify the infected individuals which are asymptomatic or pre-symptomatic, a group which is known to contribute significantly to transmission [2], [3]. By contact-tracing efforts, high-risk contacts can be identified and either quarantined or tested and quarantined to interrupt further transmission. Furthermore, the number of infected individuals detected will depend on the testing and tracing strategy and the resources available to the particular community. Wastewater-based epidemiology (WBE) is a non-invasive tool used to monitor transmission of viruses, including but not limited to fecally excreted viruses since viral shedding can involve various body fluids being discharged into wastewater systems [4]. WBE is for instance being used for the surveillance of poliovirus, the causative agent of acute flaccid paralysis [5]. In several countries, wastewater is regularly tested for the presence of poliovirus RNA in order to track the transmission of this virus in the population. Surveillance of wastewater was able to detect and characterize outbreaks of poliovirus in polio-free countries, for example in the Netherlands [6]. It is also an important tool to monitor residual poliovirus circulation in endemic countries and plays a key role in polio eradication schemes. The circulation of other enteric viruses such as norovirus, hepatitis A virus and hepatitis E virus has also been studied by wastewater surveillance [7], [8]. SARS-CoV-2 can be detected in human feces for at least three to four weeks after infection, regardless of the individual showing any symptoms or not [9]–[11]. In March 2020, the detection of SARS-CoV-2 RNA in sewage water was reported for the first time [12]. Since then, several research groups have demonstrated that SARS-CoV-2 can be successfully detected in wastewater (reviewed in [13], [14]). SARS-CoV-2 RNA remains detectable in sewage for several days to weeks, depending on conditions [15]. Surveillance of SARS-CoV-2 RNA in wastewater has been shown to be a useful tool to monitor virus circulation in the population and thus support COVID-19 mitigation strategies. Furthermore, it could serve as an early warning for SARS-CoV-2 outbreaks in a particular location and can provide valuable information on the circulation of variants of concern (VOC) or of interest (VOI) [16].

We report on our experience with SARS-CoV-2 RNA monitoring in wastewater of the city of Leuven. Leuven is the eighth largest city in Belgium with around 102.000 inhabitants. It is located in the province of Flemish Brabant in Belgium, 25 km east of Brussels. Around 50.000 tertiary education students study in Leuven’s academic institutions, the majority of which usually reside in its many student residences [17].

From September 2020 until March 2022, SARS-CoV-2 RNA was determined in municipal sewage samples, collected at a large regional sewage treatment plant. From September 2020 until May 2021, additional wastewater samples collected at local neighborhood sewers were investigated for the presence of SARS-CoV-2 RNA. Importantly, signature mutations of SARS-CoV-2 variants of concern (VOC) were observed in these wastewater samples, offering an alternative background monitoring tool for the arrival of VOC in a particular geographical area, in addition to the analysis of diagnostic patient samples. Information on sewage monitoring of specific building complexes and neighborhoods coincided with the well documented mass screening of a student residence in the drainage area of the sewage sampling site on two occasions, which allowed for temporal correlation of results from both sampling methodologies.

## MATERIAL AND METHODS

### Sampling

Wastewater was collected from various sewers in the city of Leuven. Samples were collected from four local neighborhood sewers (residentials A – D) and one larger regional wastewater treatment plant (WWTP Aquafin). Samples were taken twice a week at the following locations:

1. Residential A: residential blocks with 1295 residents
2. Residential B: area with 476 residents
3. Residential C: 313 student residences with no other residential facilities
4. Residential D: 685 residents of which 468 are students
5. Wastewater treatment plant (WWTP) Aquafin (Leuven)

At the residential sites samples were collected through a time-proportional automated sampler. For residentials A, B and C this was done using a 6712 portable sampler (Teledyne ISCO) (Figure 1). For residential D this was performed by a GLS sampler (Teledyne ISCO). The samplers were hung in a sewer manhole with a vinyl suction line hanging into the wastewater. A peristaltic pump was used to pump wastewater samples through the suction line into the sampler collection bottles. Sampling occurred between 6 and 10 AM, 100 mL samples were collected at a 15 minute interval. The 6712 collected daily samples. The GLS collected samples on Monday and Thursday.

**Figure 1:**
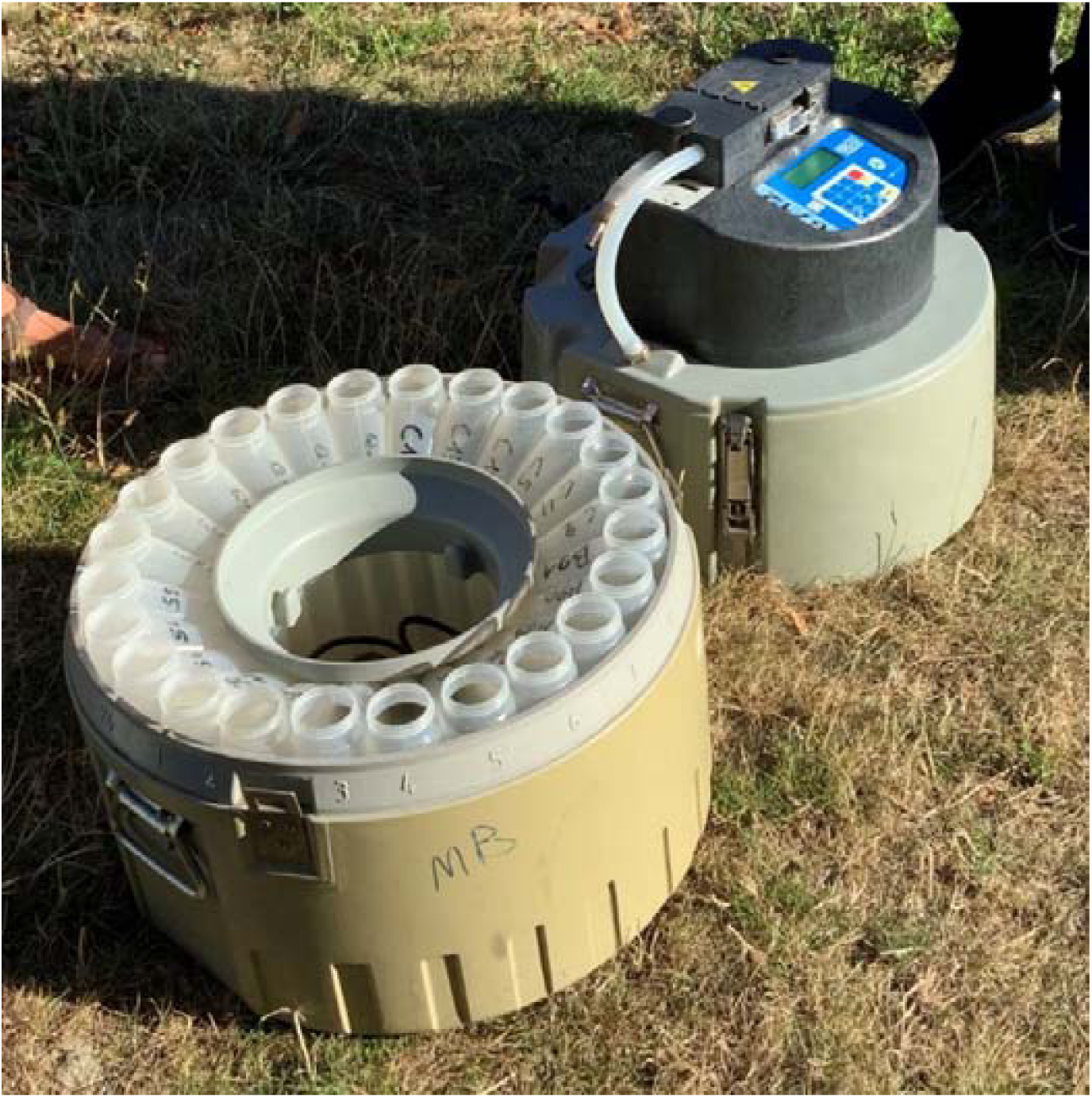
Time-proportional automated wastewater sampling device

Aquafin is a company in Flanders (Belgium) with a wastewater treatment plant (WWTP) that collects and treats municipal wastewater. The treatment plant in Leuven covers an area of approximately 115000 inhabitants. Bi-weekly samples (500 mL) of 24-hour composite influent wastewater were collected through a time-proportional automated sampler, which collects 50 mL of wastewater every 10 minutes in a large container. The samples were stored in a refrigerator at 4°C before transport to the laboratory.

### Virus concentration

Wastewater samples have high concentrations of inhibitory factors such as heavy metals, suspended solids or other organic matter. These can interfere with molecular methods for virus detection. Therefore, a pre-treatment of the raw sewage samples is necessary. First, 500 mL wastewater was centrifuged for 15 minutes at 5000g, after which 450 mL of the supernatant was transferred to an Amicon Stirred Cell device (Merck Millipore, REF UFSC40001). Using compressed air, the water is pushed through an ultrafiltration filter with a molecular weight cut-off of 30 kDa (Ultracel 30 kDa, Merck Millipore CAT PLTK07610). After all the water went through the filter, the residue and filter membrane were put in a 50 mL tube with 4,5 mL of Sigma water, resulting in a 100-fold concentrated water sample. The residue was dissolved in the water by vortexing the 50 mL tube intensely. This mixture of residue and viruses was first centrifuged for 10 minutes at 5000g and then filtered by a 1 µM syringe filter to reduce remaining inhibitors. The filtrate was used for detection of SARS-CoV-2.

### Filtration

Two different sizes of Ultracel (Amicon, Merck) for ultrafiltration were tested; 30 kDa and 100 kDa. Four wastewater samples were analyzed in duplicate and Ct values were compared (results not shown). There was no difference in results between 30 kDa and 100 kDa filters for both S and N gene, indicating that intact viruses are present in the water. The size of the Ultracel filter has no impact on duration of filtration. The 30 kDa filter was further used because of the possibility to detect other smaller viruses.

### Viral RNA extraction

#### Water samples

The QIAamp Viral RNA Mini kit (Qiagen, Ref 52906) was used for extraction of nucleic acids from 500 µL concentrated and filtered sewage water. Nucleic acids were eluted in 50 µL elution buffer.

#### Feces samples

100 mg feces was mixed with 1000 mL phosphate buffered saline (PBS) and vortexed intensely. After spinning, 140 µL was used for nucleic acid extraction with the QIAamp Viral RNA mini kit (Qiagen, Ref 52906).

### Detection of SARS-CoV-2 by RT-qPCR

To detect SARS-CoV-2 in sewage water samples or feces samples, an RT-qPCR targeting two regions of the N gene (N1 + N2) was performed on a QuantStudio 7 (ThermoFisher). A reaction mix was made by combining 5 µL of TaqMan^™^ Fast Virus 1-Step Master Mix (Applied Biosystems, CAT 4444434) with 1,5 µL of primer/probe mix from the 2019-nCoV CDC EUA kit (IDT, CAT 10006606) and 8,5 µL of RNase free water. Five µL of viral RNA was added to this mix of 15 µL, totaling 20 µL. Cycling conditions were 5 minutes at 50°C, 20 seconds 95°C followed by 45 cycles of 3 seconds at 95°C and 30 seconds at 60°C.

### Quantification of SARS-CoV-2

To quantify the amount of viral RNA in the sample, a dilution of the reference material EURM-019 (European Commission) was used. This is a synthetic single stranded RNA in buffer with an N gene copy number of 7.3 × 106 copies/µL. Seven dilutions were made and the Ct value was measured by RT-qPCR of the N gene (2019-nCoV CDC EUA kit, IDT). The measured Ct-values were converted to viral copies/µL by means of a standard curve.

Since total sewage sample preparation results in a 1000-fold concentration of the sample (450 mL sewage sample was 100X concentrated to 4,5 mL of which 500 µL was used for the RNA extraction, and eluted in 50 µL (X10)), copies/µL measured in extracted RNA equals copies/mL in the original sewage water.

### Detection of SARS-CoV-2 variants with RT-qPCR

One method for the detection of signature mutations of circulating VOCs is the use of a combination of multiplex RT-qPCR assays, targeting different specific mutations. This approach was used for wastewater samples. Even in the presence of low viral loads, mutations could still be detected and variants could be identified. Per sample, three multiplex RT-qPCR assays were performed, targeting a total of 10 signature mutations of the S gene. Primers and probes were designed by Qiagen and are shown in table 1a. Combination of signature mutations used for identification of VOC is shown in table 1b. To obtain the amplifications, a mix was prepared with TaqMan™ Fast Virus 1-Step Master Mix (Applied Biosystems, CAT 4444434), 1 µL of each primer (8 µM) and 1 µL of each probe (4 µM) and supplemented with RNase free water until a total of 15 µL is obtained. Five µL of viral RNA is added to become a total of 20 µL. Thermal cycling conditions are 5 minutes at 50°C, 2 minutes at 95°C, followed by 45 cycles of 3 seconds at 95°C and 30 seconds at 60°C using QuantStudio 7 Flex (Applied Biosystems). Analysis was performed using the QuantStudio Real-Time PCR Software.

**Table 1a:**
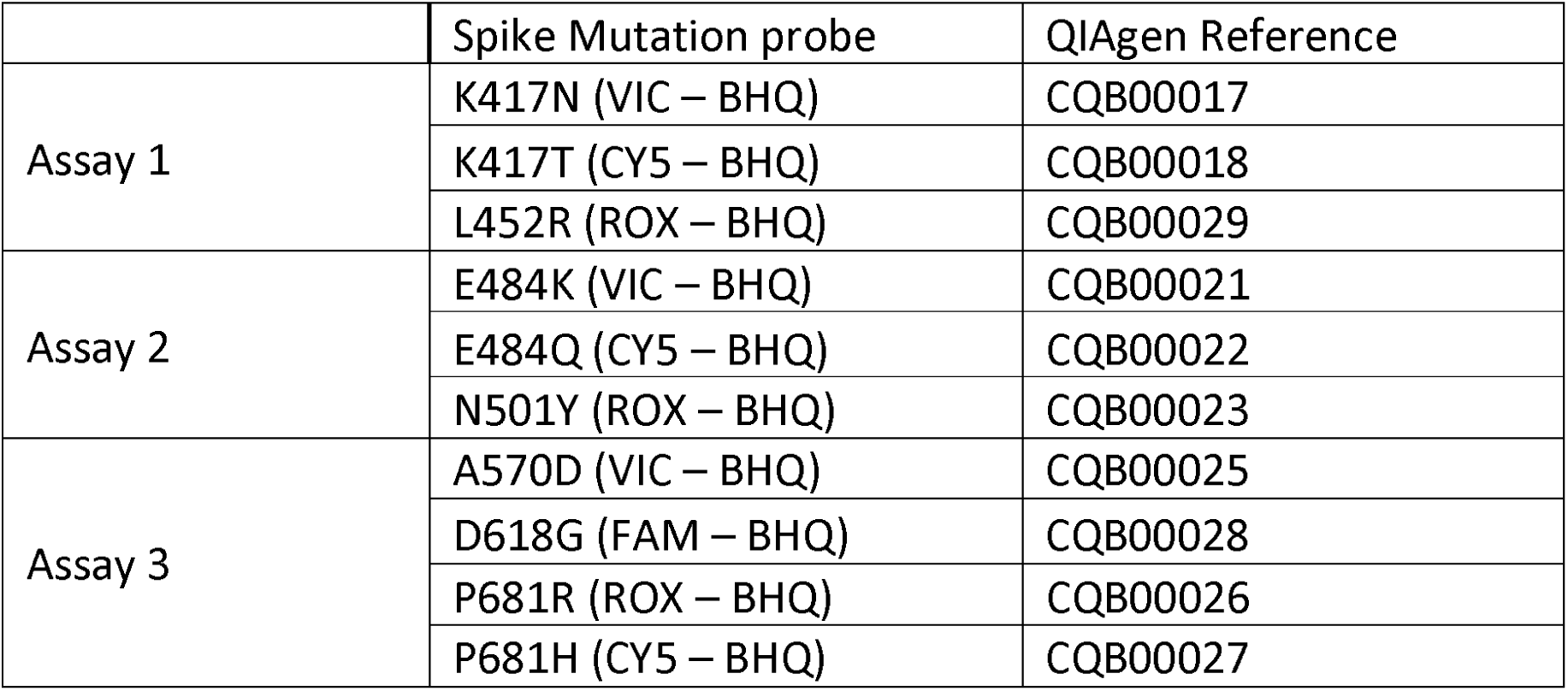
Primers and probes designed by Qiagen used for multiplex RT-qPCR targeting 10 signature mutations in the SARS-CoV-2 S gene

**Table 1b:** Signature mutations used for identification of VOC using multiplex RT-qPCR

| WHO label | Spike mutations |
| --- | --- |
| **Alpha** | N501Y<br>P681H |
| **Beta** | K417N<br>E484K<br>N501Y |
| **Gamma** | K417T<br>E484K<br>N501Y |
| **Delta** | L452R<br>P681R |
| **Omicron**<br>**BA.1 – BA.2 – BA.3** | K417N<br>N501Y<br>P681H |
| **Omicron**<br>**BA.4 – BA.5** | K417N<br>L452R<br>N501Y<br>P681H |

### Complete genome sequence of SARS-CoV-2 from a wastewater sample

Complete genome sequencing was performed by means of MinION sequencing (Oxford Nanopore Technologies, UK) methods are described in the paper of Wawina-Bokalanga et al [18].

### SARS-CoV-2 infected persons in Leuven

The daily case numbers of COVID-19 infections in Leuven were obtained from the COVID-19 dashboard of Sciensano [19]. These numbers contain all positive PCR tests from all laboratories in the greater Leuven area. COVID-19 patients were diagnosed by RT-qPCR test carried out by the laboratory of the National Reference Centre for respiratory viruses, which is part of UZ Leuven. The addresses of positive COVID-19 cases diagnosed in the testing and analysis facilities of UZ Leuven, KU Leuven and MCH and residing in the greater Leuven region were geo-coded and mapped onto the drainage areas of residential A and B.

### SARS-CoV-2 in feces samples

To assess the level of fecal virus shedding of SARS-CoV2 variants, feces was tested from patients in UZ Leuven that were infected with different variants and Ct values of respiratory samples and feces samples were compared. Twenty feces samples (leftover of SOC samples used for diagnosis) were collected from patients hospitalized in UZ Leuven. Eight feces samples from patients infected with the SARS-CoV-2 delta variant and twelve feces samples from patients infected with SARS-CoV-2 omicron variant (6 BA.1 and 6 BA.2) were screened with the N-gene RT-qPCR. Respiratory samples of the same patients were tested with the same RT-qPCR for comparison of Ct values.

Samples of patients hospitalized in UZ Leuven were used in this study according to the opting-out procedure of the hospital [20]. This states that any bodily material remaining after a diagnostic examination or procedure (residual tissue) may be used for academic research, unless the patient objects to this use. The use of patients residual materials by the National Reference Center for coronaviruses for SARS-CoV-2 research was approved by the UZ Leuven Ethics Committee.

## RESULTS

### Wastewater surveillance in different residential areas of Leuven

Sewage samples from different local sewers of smaller residential areas in Leuven were collected, starting from September 2020 to June 2021, and compared to the number of positive cases reported for these areas.

In the wastewater of the Residential A, representing a total of 1295 residents, the first uptick of the SARS-CoV-2 viral load was detected on September 27^th^ of 2020 (Figure 2). Positive cases followed rapidly and peaked in October 2020 with 5 detected positive cases over a period of 3 weeks. No wastewater samples were taken during the Christmas holiday of 2020/2021, so it cannot be established whether the rise in positive cases in January 2021 was preceded by an increase in wastewater viral load or not. However, the viral load peak of SARS-CoV-2 in residential A wastewater on March 31^st^ of 2021 was again followed by a rise in positive cases among the residents (with a peak of 18 cases/3 weeks at the end of April 2021),

**Figure 2:**
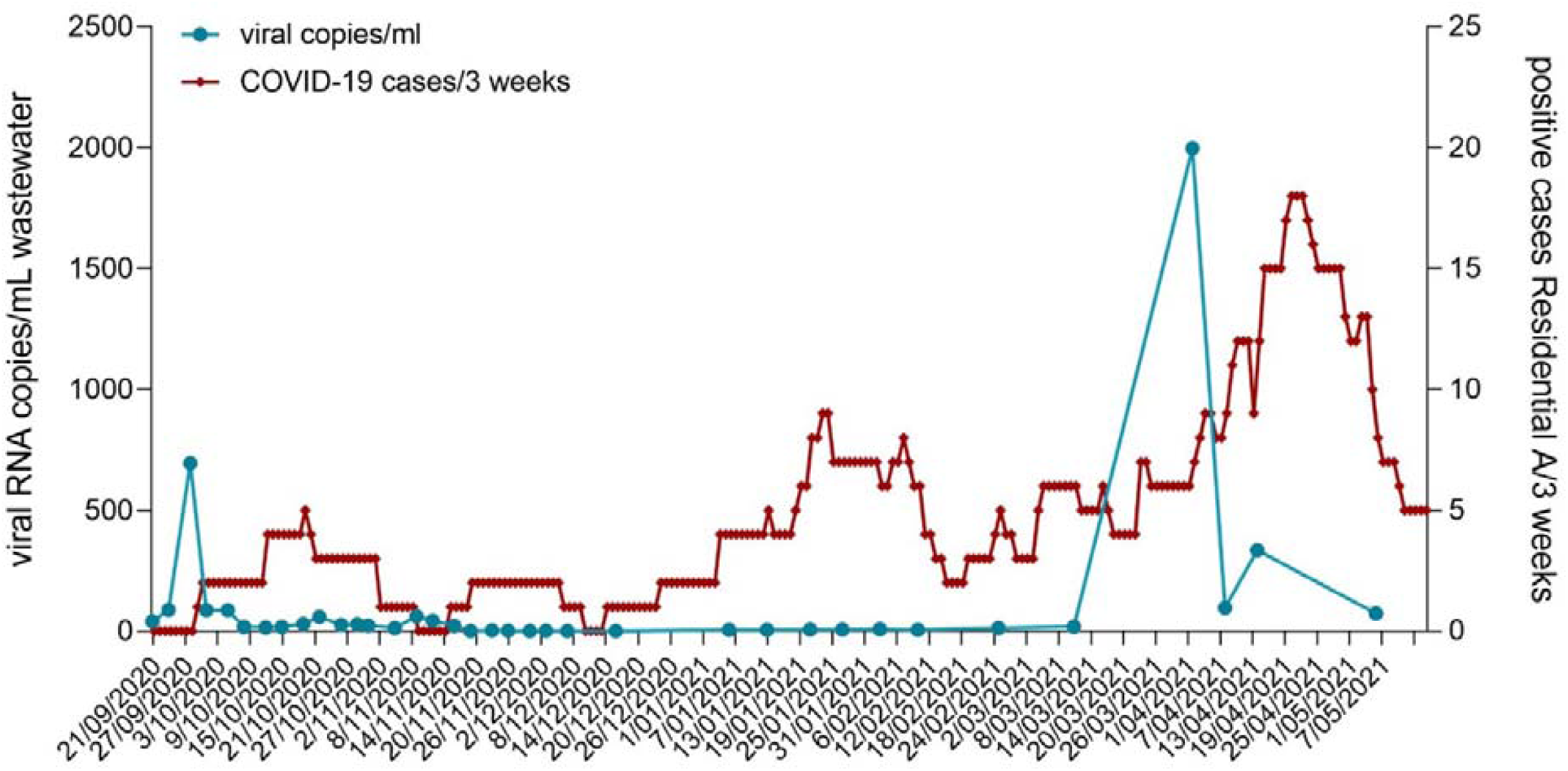
Correlation between the number of positive cases in Residential A (per 3 weeks) and the viral RNA levels found in wastewater of this Residential

In residential B, with 476 residents, wastewater was collected from October 2020 to May 2021 (Figure 3). Also in this district, there were two early warning peaks in the wastewater viral load, where a rise of viral copies/mL SARS-CoV-2 was seen before the number of positive cases went up, in October 2020 and in January 2021. In March 2021 the concentration of virus was again elevated in the wastewater, but surprisingly the number of positive cases did not follow, even though the 3^rd^ SARS-CoV-2 wave had started at that moment. An explanation might be that residents mainly experienced asymptomatic infections, were not tested and were hence not registered by Sciensano.

**Figure 3:**
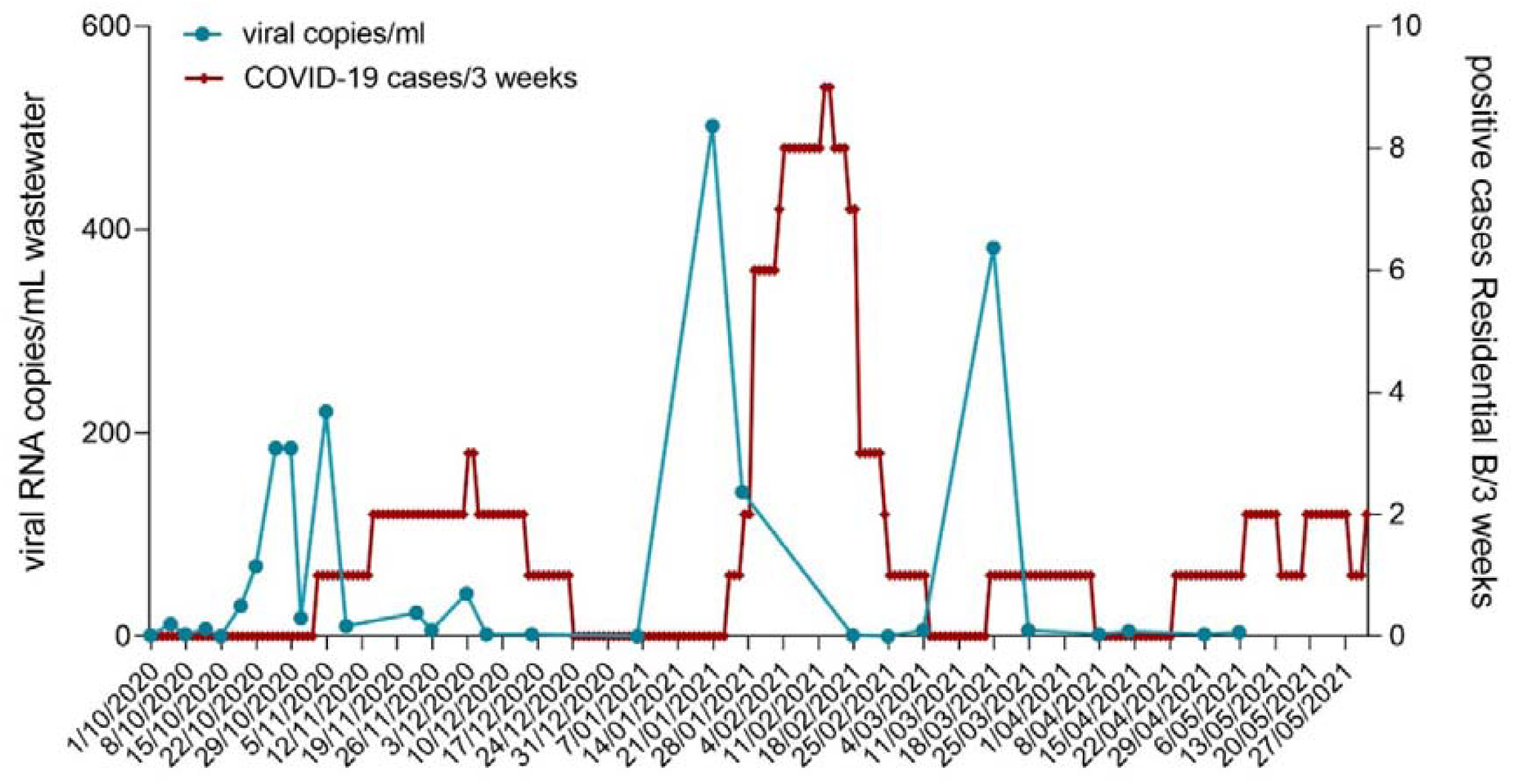
Correlation between the number of positive cases in Residential B (per 3 weeks) and the viral RNA levels found in wastewater of this Residential

For residential C and D the data was not complete due to some technical problems with the sampler or blockage of the sampler. Therefore, no figure was created with results for these 2 residentials. However, using the available samples from residentials C and D we were able to provide useful information to support contact tracing and screening.

### Tracing of asymptomatic infected persons based on wastewater surveillance

The university of Leuven has implemented a comprehensive contact tracing approach for its 50.000 students since October 2020. On two occasions, a sudden increase in viral RNA in sewage of a particular residential area concurred with the screening of a large student residence in the testing site’s drainage area, allowing us to correlate both the temporal dynamics of sewage and biological sampling as well as the possibility of linking VOC identification and WGS in sewage samples to their identification in individuals living in the upstream residential area. On one occasion, the uptick of SARS-CoV-2 initiated the large-scale screening of the upstream residence. In another, screening of a student residence was ongoing when the contact tracing team was informed about the uptick in viral load in the sewage sampling site draining the outbreak location.

### Reactive screening of a student residence based on sewage water

In the first case, a student residence just outside of the city center of Leuven was concerned. The sewage sampling site of residential C drains no other buildings than this complex and is not mixed with rainwater. The complex contains 261 rooms, 16 studio’s and 14 apartments. Kitchen and sanitary facilities are shared by 12 to 13 students sharing a living unit.

A case of COVID-19 was detected in the complex at the end of March by qPCR test. Ct value was 25.4 for the N gene. The S gene was not detected, indicating the presence of the Alpha variant [21], [22].. In the beginning of April, the KU Leuven contact tracing team was alerted to the fact that there was a rise in concentration of SARS-CoV-2 in the wastewater samples of the student’s residence. On March 18^th^, the wastewater was still negative but on March 25^th^ and April 2^nd^ Ct values of 27,1 and 27, 3 were measured. Since a recent index case was known in the building, it was decided to screen the whole facility since the index case could not be reached and queried on whether they had been in isolation in the student residence or not.

Of the 269 current residents of the facility, 62 reported not having recently resided in the building. They were deemed non-eligible for testing. Of the 207 remaining residents, 188 were reached by the contact tracing team. 19 were lost to follow-up. While 22 students refused testing, 143 underwent screening by a nasopharyngeal sample and qPCR at least once.

One additional COVID-19 case was identified. This individual was asymptomatic and was diagnosed after screening. Ct value was 29.5 for N gene, while the S gene was not detected. They had no alternative source of infection than the student residence.

WGS was performed on the clinical sample of the index case and the wastewater sample of March 25^th^. Both strains were exactly the same and resulted in a B.1.1.7 or Alpha variant with specific mutations and deletions in the S gene: H69-/ V70-/Y144-/ N501Y / A570D / D614G / P681H / S704L / T716I / S982A / D1118H / K1205N.

### Concurrence of an uptick of SARS-CoV-2 in sewage with an ongoing screening effort of a large student residence in the sampling site’s upstream residential area

In a second case, a student residence located in Residential D was experiencing a large outbreak of COVID-19 when the contact tracing team was alerted about an uptick in the SARS-CoV-2 concentration of the sewage system draining the residential area comprised of 685 residents including 468 students. The complex contains 405 rooms and 26 studios. Each room has their own sanitary facilities. A kitchen is shared by 10 to 18 students of the same living unit. A first case was diagnosed in the beginning of April 2020. Since several secondary cases were identified, it was decided to invite all residents having recently resided in the residence for testing.

On April 2^nd^ the wastewater sample was negative. On April 8, unfortunately there was no wastewater sample of this residence due to a blockage of toilet paper in the sampling device. On April 14, the Ct value of the wastewater was 26.6 (608 copies/mL). All living units were placed into quarantine until screening results were available for the unit. If at least one index-case was identified within the unit, quarantine was extended and all students within the unit were screened every 7^th^ day until no additional secondary cases were identified, at which point quarantine was lifted for the particular unit.

Of the 468 residents, 143 had not recently resided in the student residence and 39 were deemed not at risk by the contact tracing team and therefore excluded from screening. Of the 287 students requiring testing, 10 were not reached and 7 refused testing. The remaining 269 students underwent at least one diagnostic test in the weeks following the identification of the first index case. Between April 9^th^ and 28^th^ of, 24 cases of COVID-19 were registered (positivity rate of 9%), of which 23 were epidemiologically linked to the student residence based on contact tracing information. In all samples for which typing by VOC PCR and/or WGS was possible, the Alpha variant (Pango lineage B.1.1.7) was detected. This is in accordance with the epidemiologic situation in Belgium during that period, with Alpha being the dominant variant, representing between 80 and 85% of circulating strains [23].

### Detection of SARS-CoV-2 variants in the wastewater treatment plant (WWTP) of Leuven

Municipal wastewater samples of the city of Leuven were collected at the sewage treatment plant Aquafin starting from September 2020 until March 2022. Aquafin collects household wastewater from the municipal sewers in collector sewers. As expected, viral RNA of SARS-CoV-2 was detected in the city wastewater samples at all time points. The viral RNA levels in the city sewage mostly followed the same trend as the number of positive cases in Leuven per 3 weeks (Figure 4). During the second coronavirus wave, although the number of positive human cases in Leuven started to increase on October 5^th^ 2020, there was only a slight increase in sewage viral RNA as of October 12^th^. A more pronounced increase was observed on October 19^th^. During the third wave the viral copies in wastewater increased in April 2021, but there was no increase in positive cases at that time. This might be due to a high proportion of infections being asymptomatic, which remained undetected. In July 2021 and September 2021 there were also upticks in the wastewater without an associated increase in positive cases in Leuven. In November 2021, the Delta variant caused a fourth wave of infections. The concentration SARS-CoV-2 in wastewater was rising fast and correlated well with the increase of positive cases. Remarkably, during the start of the fifth wave which was caused by Omicron BA.1 infection, the number of positive cases went up very fast but the concentration of the virus in wastewater remained low, and a correlation between both was no longer observed. The wastewater monitoring strategy that was used in Leuven was thus unable to provide early warnings of an increase in the number of positive cases in the community. Since the emergence of Omicron BA.2, viral loads in wastewater increased again as seen in Figure 4, and the correlation between infected patients and viral load in wastewater could be observed again.

**Figure 4:**
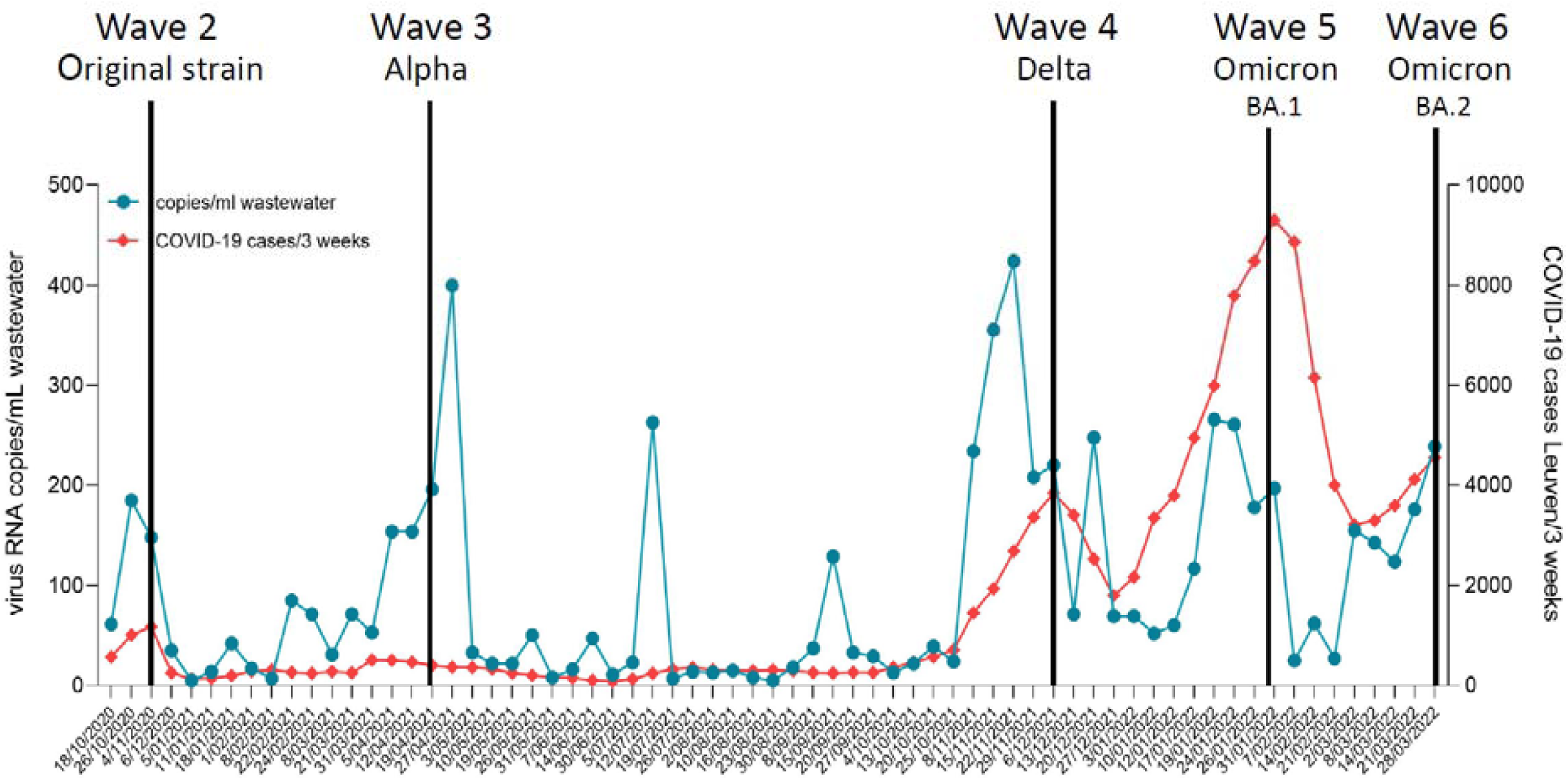
Correlation between the number of positive cases in Leuven (per 3 weeks) and viral RNA levels in wastewater from the Aquafin WWTP in Leuven.

Using a combined multiplex qPCR, VOCs of SARS-CoV-2 could be distinguished. The SARS-CoV-2 RNA in wastewater samples collected in December 2020 did not contain any signature mutation, indicating the wild type (Wuhan) strain was present at that time. From January 18^th^ 2021 the mutations N501Y and A570D were detected, which are indicative of the Alpha variant. From April 2021 on, the E484K mutation was sporadically detected. This mutation is present in the Beta and Gamma variants. In June 2021 the L452R mutation, a signature mutation for Delta, was first detected in the wastewater of Aquafin Leuven. At that time, the Alpha variant was also still detected in the wastewater, and co-circulated with Delta until July 2021. Delta was detected until January 2022. The K417N mutation, indicative of Omicron, was first seen in a sample dating from December 27^th^ 2021. These findings are shown in Figure 5. Using our multiplex RT-qPCR assay, we were unable to distinguish between Omicron BA.1 and BA.2. The circulating variants in the WWTP of Leuven are in accordance with the VOC data published by the Belgian National reference lab (figure 6).

**Figure 5:**
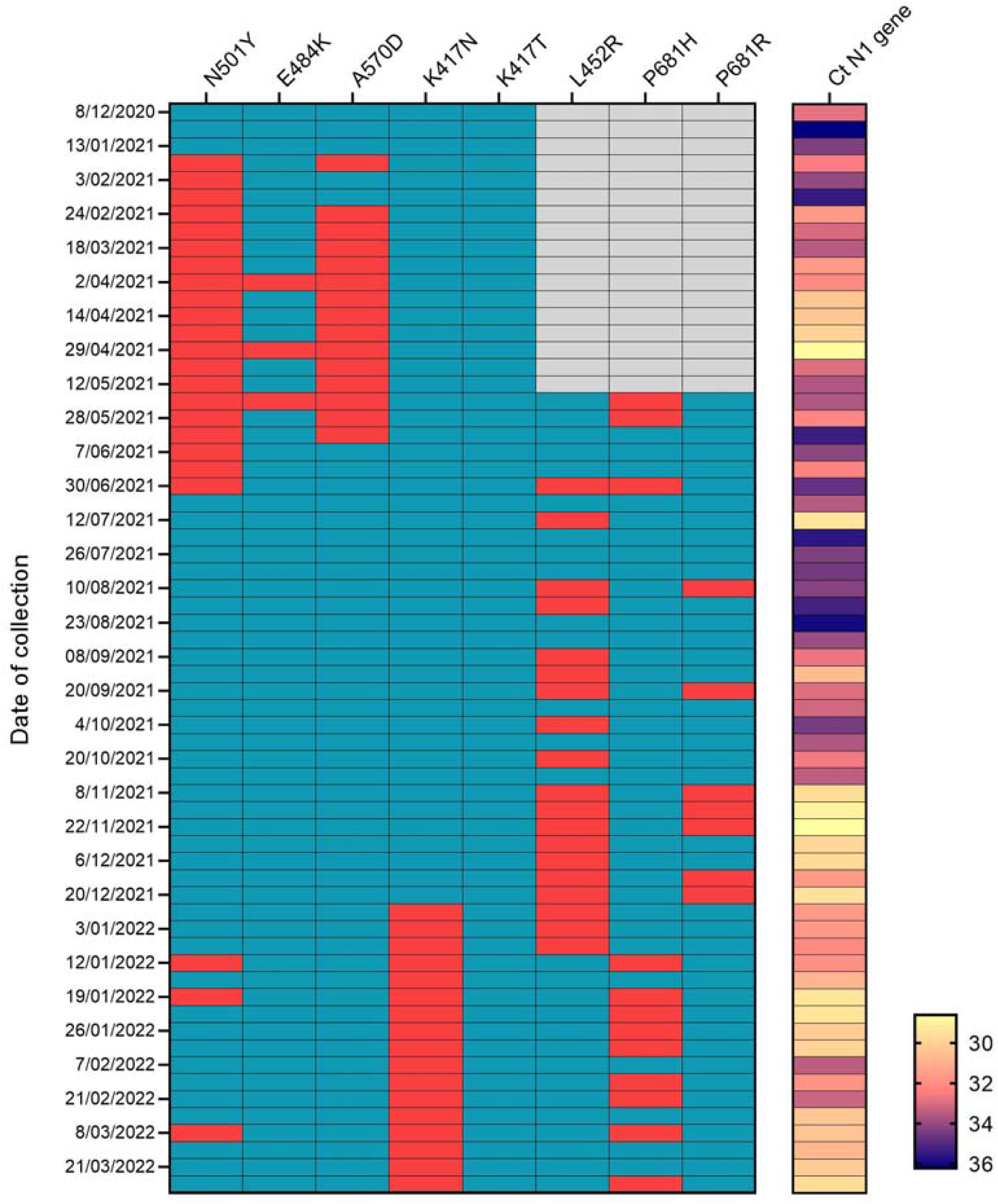
SARS-CoV-2 signature mutations detected in weekly wastewater samples from WWTP of Leuven.

**Figure 6:**
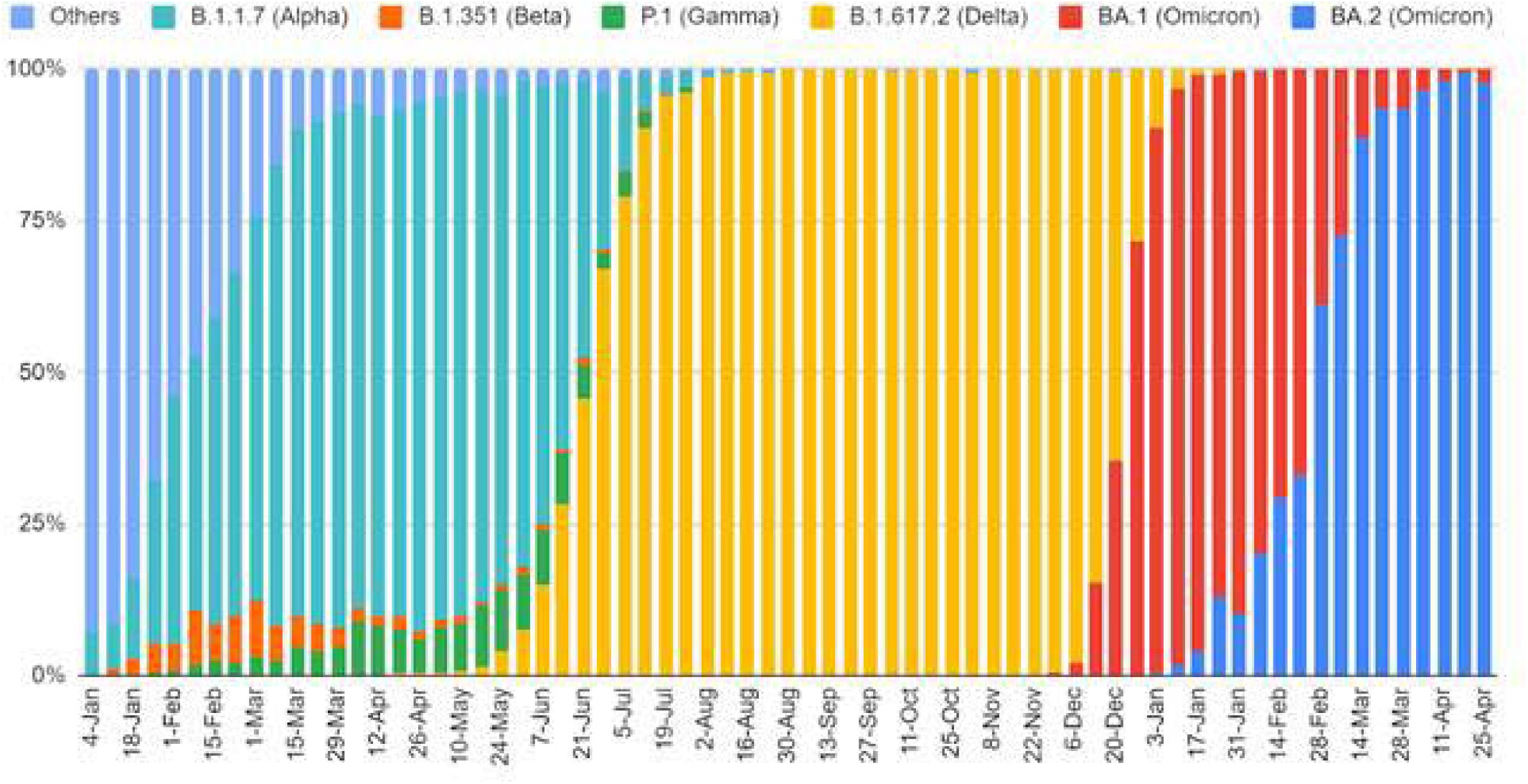
Share of variants of concern per week in Belgium, from January 2021 until April 2022

### Excretion of Delta and Omicron in fecal samples

Results of excretion in feces of Delta and Omicron positive patients were presented in figure 7. Whereas the Delta and Omicron BA.2 curve shows an irregular curve with lower Ct values representing higher viral loads, the Omicron BA.1 excretion shows a steady line, around Ct 30. These results indicate that that the fecal excretion of Omicron BA.1 is overall at a low level, whereas Delta excretion and Omicron BA.2 can be much higher, which could explain why the large wave of Omicron BA.1 cases was not accompanied by an equivalent viral load peak in wastewater (Figure 4). When BA.2 was more represented in the positive cases, the viral load in wastewater was rising again.

**Figure 7:**
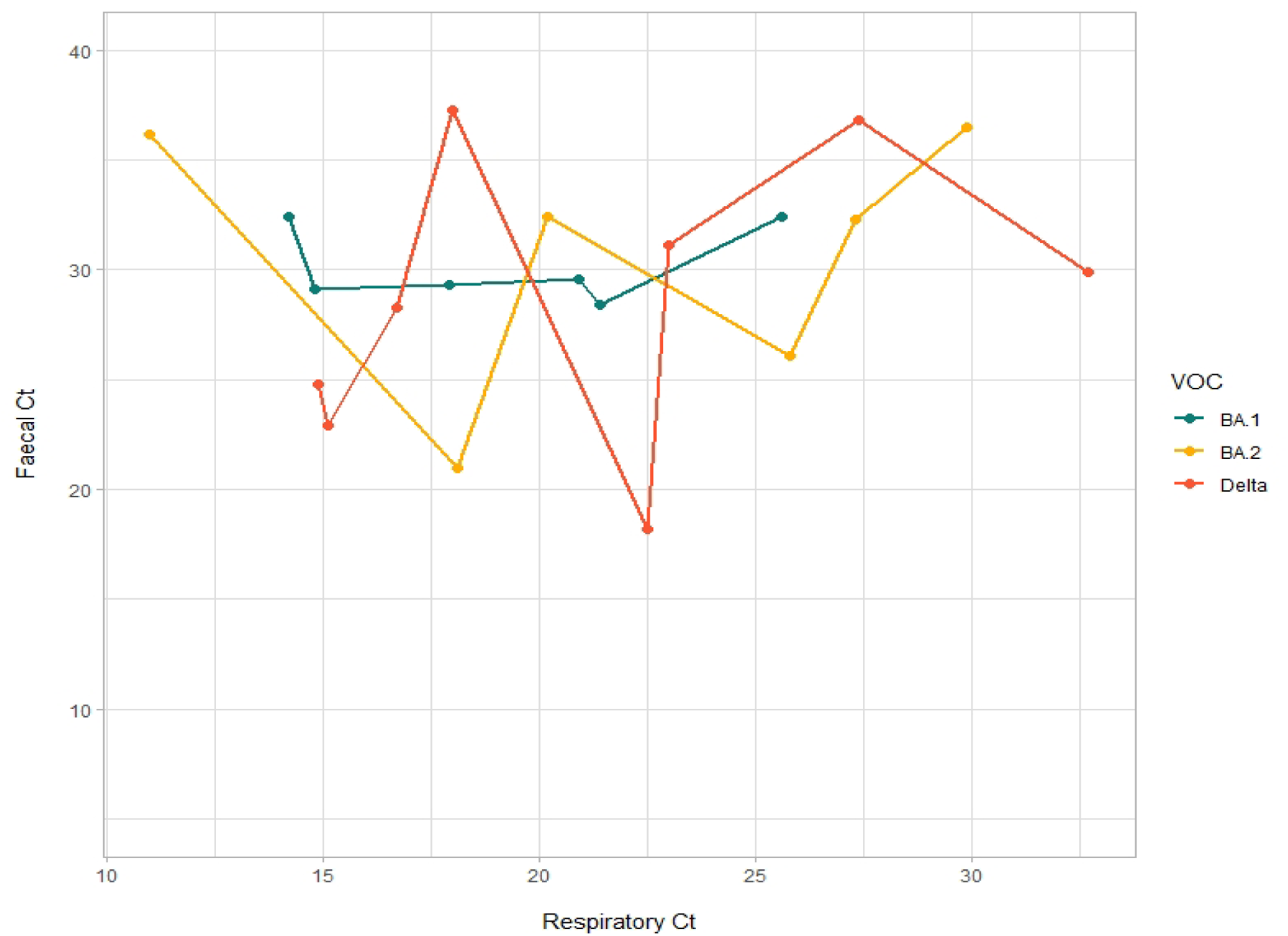
Difference of viral RNA levels SARS-CoV-2 found in feces of infected patient

## DISCUSSION

Wastewater surveillance is a rapidly developing research area and is already playing an important role in the management of the COVID-19 pandemic all over the world. We developed a pilot project of wastewater surveillance in Leuven to investigate whether it can be used as an early warning system. The wastewater of four different neighborhoods in Leuven was tested for SARS-CoV-2 during 8 months and one WWTP was screened for 16 months. The results of residentials A and B show a peak of viral SARS-CoV-2 copies before the peak of positive cases, indicating that wastewater surveillance can indeed be used as an early warning system. If COVID testing of individual patients will be further reduced in the future, wastewater testing can detect an outbreak on a building level, making it a very valuable tool in times of endemics or pandemics. This applies not only to SARS-CoV-2, but to several other pathogens as well as studies related to pharma and drug consumption [24].

In wastewater of neighborhoods and districts the early warning signal was very clear. Also, the viral copies measured were much higher in these places than WWTP because it belongs to a smaller community, and sewage water is not always mixed up with rainwater. A possible dilution caused by inflow of rainwater can be calculated by measuring concentration of pepper mild mottle virus (PMMoV) in the sample and normalizing the SARS-CoV-2 gene copies to PMMeV gene copies. PMMeV is a fecal biomarker present in human feces through diet [25], [26]. For this pilot project, we did not take possible rainwater infiltration into account. For WBE and wastewater surveillance this should be implemented [27]. Two occasions were documented where an uptick in SARS-CoV-2 concentration in the sewage system was temporally correlated with a large-scale screening effort of the whole or the large majority of the upstream human population in a residential setting. On one occasion, the uptick in SARS-CoV-2 in sewage even initiated the screening effort. We have demonstrated a tight temporal fit between an increased infection rate of SARS-CoV-2 and an increased detection in sewage, although this temporal fit depends heavily on the frequency of sampling of the sewage system, the barrier to testing for the upstream human population and the speed and comprehensiveness of the contact tracing efforts in the area. In both instances recorded in the current paper, the first positive sewage sample corresponded exactly with the first SARS-CoV-2 diagnosis in the upstream residential complex. We also show the sewage signal to be highly sensitive, having picked up two ongoing infections in just over 250 inhabitants. In all analyses performed (city wide level, neighborhood level), there appears to be a ‘dose response’ relationship between the sewage sampling and the upstream SARS-CoV-2 transmission, with varying delays depending on the frequency of sampling and the size of the upstream drainage area. Additionally, we have shown that it is feasible to organize background genomic surveillance by using the ‘collective sample’ of humans in a particular area by typing the circulating variants in the sewage system.

In the WWTP (Aquafin), the early warning effect was less visible. Nevertheless, most peaks of viral copies were in correlation with peaks of positive cases, which is also reported by other research groups around the globe [28], [29]. We were able to detect different SARS-CoV-2 variants during the different waves with specific key mutations starting from N501Y for the Alpha variant and ending with K417N for the Omicron variant. The transition of variants was noticeable in the wastewater, with key mutations of two different variants cocirculating during the period of a shift in variants in the population. In the future, we also want to focus on the detection of novel SARS-CoV-2 variants in wastewater samples by whole genome sequencing. In this perspective, biobanking of the samples that were collected over more than one year has been done to allow retrospective analysis, and can provide crucial information about circulating virus variants.

Peaks of viral loads in the wastewater were seen simultaneously with the waves of the original Wuhan SARS-CoV-2 strain, the Alpha variant and the Delta variant. With the Omicron variant however, the viral load peak in wastewater was not that clear: the increase of virus in wastewater came much later than the increase in positive cases and did not reach a high viral load despite a high concurrent incidence rate. The reason why Omicron BA.1 was not easy to detect in wastewater was therefore further investigated. Firstly, the primer-probe sets were aligned against both Omicron BA.1 and BA.2. De N1 probe of the CDC primer/probe kit (2019-nCoV CDC EUA kit) has only one nucleotide that is mismatching with Omicron. Normally this would not affect test results [30]. The N2 primer-probe set was in complete alignment with the BA.1 and BA.2 sequences. To make sure the N1 probe is not the reason for a decreased sensitivity in detection of Omicron BA.1, different primer-probe sets were used, resulting in comparable Ct values (data not shown). This indicated that the primer/probe set is not responsible for the lack of correlation of Omicron infections and viral loads in wastewater [31]. Secondly, fecal samples from SARS-CoV-2 positive patients, infected with different variants, were collected to verify a possible difference in fecal excretion of virus. Results in figure 7 show a difference in fecal excretion between Delta, Omicron BA.1 and Omicron BA.2. The viral loads in feces of Omicron BA.1-infected patients were not as high as the viral load in Delta or Omicron BA.2 infected patients. We hypothesize that the absence of an observable correlation between the number of infections with Omicron BA.1 and viral loads in wastewater samples, is due to a lower excretion of Omicron BA.1 via feces compared to other variants. The reason behind this needs further research.

## CONCLUSION

Our study in Leuven showed that the objectives stipulated in the WHO Guidance on environmental surveillance for SARS-CoV-2 to complement public health surveillance [16], namely to provide early warning and additional evidence regarding the virus circulation in the population, including its presence or absence, trends in concentrations, and presence of VOCs, could be met with our set-up. Tracking increasing and decreasing trends at community level to help target COVID-19 response and interventions was done on the city level by tracking four different sites in the city of Leuven. At two sites, increasing viral loads triggered an intervention by the contact tracing team. Detection of local outbreaks can be done at any place were the sampler can be attached into a sewer system. The identification of VOCs was done by the VOC qPCR of Qiagen that detected circulating variants in the WWTP of Leuven in accordance with the VOC data published by the Belgian National reference lab. All SARS-CoV-2 variants could be detected in untreated wastewater. There were however marked differences in viral loads, with the Omicron BA.1 wave corresponding to lower levels of detectable SARS-CoV-2 RNA in wastewater compared to other variants. The question why some variants are less detectable was solved by screening fecal samples, with viral loads of BA.1 being lower that other VOCs detected in feces.

Wastewater surveillance can be an important objective indicator of virus circulation in the community, and in this way complement the public health surveillance based on screening of individual patients, especially in the case of asymptomatic infections and in times of reduced test capacity or lower willingness to test.

## Data Availability

All data produced in the present study are available upon reasonable request to the authors

## ACKNOWLEDGEMENTS

We would like to thank the technical service of the city of Leuven and the technical service of UZ and KU Leuven for their assistance and hard work to collect all the samples in this study.

Special thanks to: Jan De Witte, Joris Doucet, Stef De Ridder, Sam Nagels and Gijs Van Pottelbergh.

